# End-to-End Deep Learning for Detecting Metastatic Breast Cancer in Axillary Lymph Node from Digital Pathology Images

**DOI:** 10.1101/2021.04.09.21255183

**Authors:** Turki Turki, Anmar Al-Sharif, Y-h. Taguchi

## Abstract

Metastatic breast cancer is one of the attributed leading causes of women deaths worldwide. Accurate diagnosis to the spread of breast cancer to axillary lymph nodes (ALNs) is done by breast pathologist, utilizing the microscope to inspect and then providing the biopsy report. Because such a diagnosis process requires special expertise, there is a need for artificial intelligence-based tools to assist breast pathologists to automatically detect breast cancer metastases. This study aims to detect breast cancer metastasized to ALN with end-to-end deep learning (DL). Also, we utilize several DL architectures, including DenseNet121, ResNet50, VGG16, Xception as well as a customized lightweight convolutional neural network. We evaluate the DL models on NVIDIA GeForce RTX 2080Ti GPU using 114 processed microscopic images pertaining to ALN metastases in breast cancer patients. Compared to all DL models employed in this study, experimental results show that DenseNet121 generates the highest performance results (64– 68%) based on AUC and accuracy.

## I. Introduction

Breast cancer metastasis is associated with increased mortality rates attributed to the possible spread of cancer cells outside the tumor through lymphatic vessel to other areas in the lymph system, including lymph nodes in axilla, near collarbone and breast (see Figure 1) [1-3]. To accurately diagnose breast cancer metastasized to lymph nodes, a biopsy specimen is obtained from the patient and is provided to a breast pathologist, using a microscope associated with camera to inspect the specimen via a computer software through manual whole slide imaging (see Figure 2). Such a diagnosis process requires experience and is time consuming when dealing with many patients [4-7].

**Figure 1:**
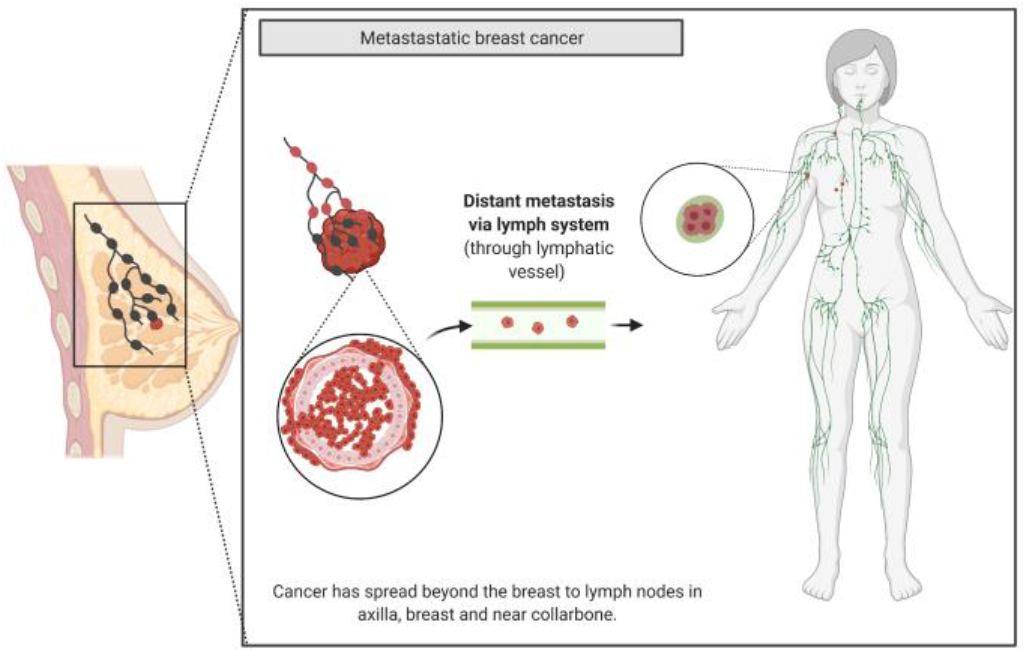
An overview showing breast cancer metastasized to several lymph nodes. Figure created with Biorender.com.

**Figure 2:**
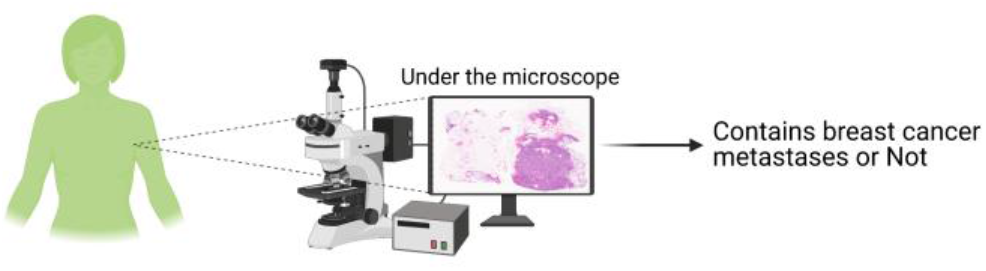
Whole slide imaging is done by a pathologist in which a biopsy with H&E stain is placed under the microscope, including an attached camera. Figure created with Biorender.com.

Accurate and early detection of breast cancer metastases can help pathologists to avoid examining breast cancer under the microscope and rapidly reporting the status of breast cancer patients. Therefore, patients can immediately start the treatment plan with medical oncologists and surgeons. Because the treatment plan provided by oncologists and surgeons depends on the diagnosis and report obtained from pathologists, researchers have proposed computational methods using artificial intelligence to assist pathologists. For example, Lee et al. [8] utilized convolutional neural networks to predict breast cancer metastases to axillary lymph nodes (ALNs). They utilized DenseNet-121 to build a deep learning (DL) model, which was then compared against other machine learning algorithms (including logistic regression, SVM, and XGBoost) with handcrafted features. Experimental results demonstrated that DenseNet-121 performed better than machine learning algorithms.

Zhou et al. [9] proposed a DL approach for the automatic prediction of lymph node metastasis in breast cancer patients, working as follows. A training and validation sets consisting of 877 images, where 441 images belong to patients with lymph-node metastases while 436 images are for patients with no lymph-node metastases. Several DL architectures were adapted and utilized for this binary classification tasks, including Inception-ResNet V2, ResNet-101, and Inception V3. The resulted models were evaluated using internal and external test sets (labeled by 5 radiologists), consisting of 97 and 81 images, respectively. Reported results show that Inception V3 generated the highest performance results. Zheng et al. [10] proposed a DL approach incorporating breast cancer images and clinical parameters (obtained from clinical texts such as tumor type, age estrogen receptor (ER) status and others characteristics pertaining to tumor and patients) to assist clinical oncologists in deciding if there is a need or not to perform ALN dissection or sentinel lymph node biopsy of patients. Therefore, avoiding such an unnecessary surgery and associated complications. Experimental results demonstrated the good performance results of the presented DL approach when is used with ResNet50 DL. Other researchers have proposed other DL approaches for various learning tasks pertaining to breast cancer metastases [11-19].

Although previous aim to provide computational methods to detect breast cancer metastasized to lymph nodes, this study is unique in several respects. First, we aim to utilize and adapt additional DL architectures including a customized convolutional neural network. Examples of such DL architectures include DenseNet121, ResNet50, VGG16 and Xception. Second, we process 114 gigapixel images pertaining to ALN metastases on NVIDIA GeForce RTX 2080Ti GPU. Then, resizing each image to 256 * 256 pixels to be used in DL experiments. We include these images in supplementary material. Third, we evaluate and report performance differences among DL models on the whole image dataset using five-fold cross-validation. Fourth, our reported results on NVIDIA GeForce RTX 2080Ti GPU show that DensNet121 outperformed other DL models including a convolutional neural network consisting of few layers on manual whole slide images pertaining to ALN metastases. Fifth, the presented study provides a pathological process automation using DL as shown in Figure 3.

**Figure 3:**
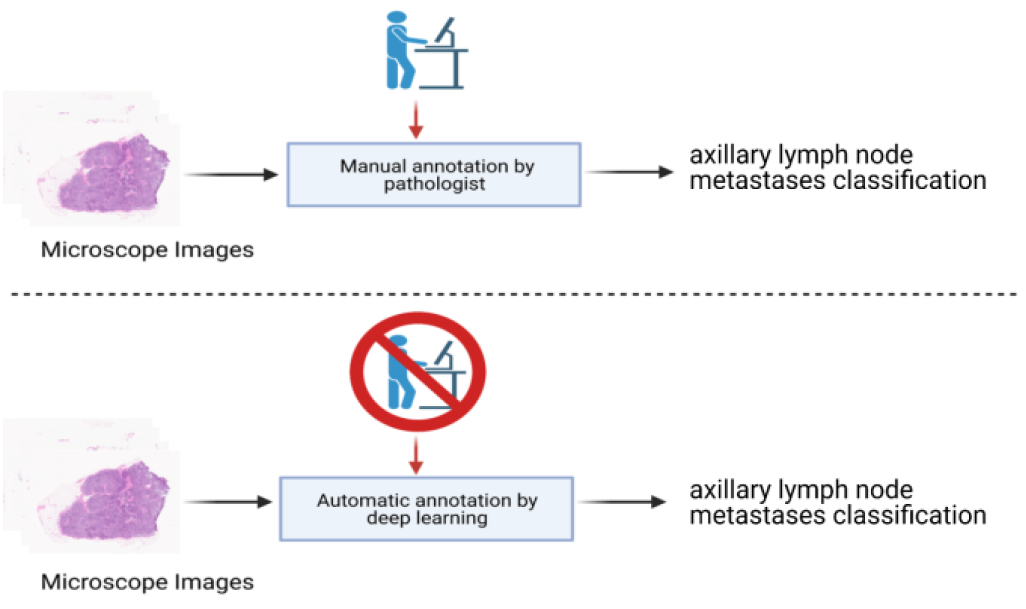
The upper panel demonstrates a manual annotation by a pathologist to determine the outcome (i.e., axillary lymph node metastases (ALN) or no ALN metastases) of microscope images in breast cancer patients. The lower panel displays an automatic annotation by deep learning (DL) technology. Figure created with Biorender.com.

The remaining of this study is organized as follows. Section II describes the image dataset pertaining to ALN metastases, as well as presenting the DL approach to address the binary classification task. Section III reports performance results related to predicting ALN metastasis. Section IV presents a discussion of the results in this study. Section V concludes the study and points out future research directions.

## II. Methods

### A. Axillary Lymph Node Metastases Image Dataset

Microscopic images for ALN metastases in breast cancer patients were downloaded from Cancer Imaging Archive at https://wiki.cancerimagingarchive.net/display/Public/Breast+Metastases+to+Axillary+Lymph+Nodes [15, 20, 21]. The dataset used in our study consisted of 114 whole slide images, where 84 had no ALN metastases and 30 images had ALN metastases. We used python script on NVIDIA GeForce RTX 2080Ti GPU to process the 114 gigapixel whole slide images, then resize them to (256 * 256 pixels). We include these processed images in supplementary material and include some pictures in Figures throughout the manuscript. It is worth noting that the original image dataset had 130 images. However, 16 images out of 130 had formatting issues. Therefore, we excluded them.

### B. Deep Learning Approach

In Figure 4, we show the DL approach used in this study to discriminate between positive and negative ALN in breast cancer patients. For a given training set 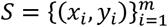 composed of *m* labeled ALN images obtained from a pathologist. If a training example is associated with class label of 1, then that indicates the presence of cancer in ALN. On the other hand, if a training example is associated with a class label of 0, then cancer is not present in the ALN. We adapted several pre-trained DL architectures, including DensNet121. ResNet50, VGG16, Xception and a lightweight convolutional neural network (named LCNN). For the pre-trained DL models, we replaced the dense layers in the densely connected classifier of each pre-trained model by two dense layers. The first dense layer includes a ReLu activation, followed by a dropout. Because we have a binary classification task, the second dense layer had 1 unit and a sigmoid activation. The LCNN had 4 layers as two pairs of interleaved Convolutional and MaxPooling layers for extracting useful features. Then, features are flattened, and the outcome is provided to a densely connected classifier consisting of three dense layers, where the last layer has 1 unit and sigmoid activation as in the adapted pre-trained DL models. For all DL models, we trained each DL model on a balanced training set of ALN images by undersampling from the majority class examples. In this study, majority class examples correspond to images of negative ALN. Then, we applied the trained models to unseen ALN images to generate predictions correspond to probabilities. If predicted probability is greater than 0.5, then it is mapped to 1; otherwise, it is mapped to 0. Prediction of 1 (i.e., considered positive) means ALNs have cancer while 0 (i.e., considered negative) means ALNs are free of cancer.

**Figure 4:**
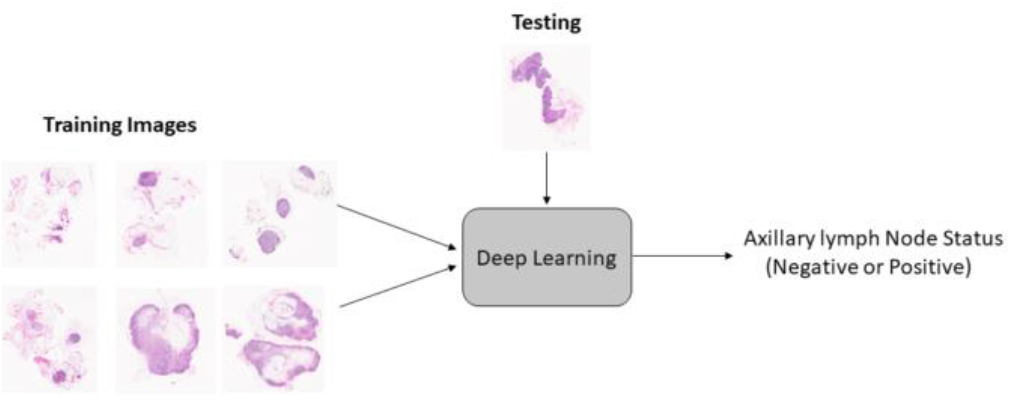
A demonstration of the deep learning (DL) approach employed in this study for discrimination between positive axillary lymph node (ALN) and negative ALN in metastasized breast cancer.

## III. RESULTS

### A. Classification methodology

We adapted five DL architectures—LCNN, DensNet121. ResNet50, VGG16, Xception—for predicting ALN metastases in breast cancer patients. All architectures work under the supervised learning scenario. That is, a training set is provided to train each DL model. Then, a test set is provided to each trained DL model to generate predictions, mapped to 1 (i.e., positive ALN) or 0 (i.e., negative ALN). We employed four performance measures: Accuracy (ACC), F1-score (F1), area under curve (AUC) and Matthews correlation coefficient (MCC). Also, we utilized five-fold cross-validation as in [22].

### B. Implementation Details

We utilized Anaconda Python 3.7.4 (with Jupyter Notebook), Keras and Sklearn libraries to run and evaluate DL experiments. For the hardware, we used NVIDIA RTX 2080Ti GPU consisting of 4352 CUDA cores with 11 GB memory.

### C. Classification Results

Figure 5 displays the combined confusion matrices of 5 test splits during the cross-validation process. If numbers in each confusion matrix is added, then we get 114, corresponding to the whole image dataset in this study. DenseNet121 generates the highest accurate predictions by accurately predicting 78 images out of 114 images. VGG16 is the second by accurately predicting 68 images out of 114 images. ResNet50 is the worst by accurately predicting 59 images out of 114. Table 1 reports performance results using four performance measures during five-fold cross-validation. It can be seen from Table 1 that DenseNet121 generates the highest performance results, shown in bold. These results demonstrate the good performance of DenseNet121.

**Table 1:** Performance comparison between different deep learning (DL) models (during 5-fold cross-validation) for predicting positive axillary lymph nodes (ALNs) and negative ALNs. MACC is the mean accuracy. MF1 is the mean F1. MMCC is the mean MCC. SD is the standard deviation. The highest performance result is shown in bold.

|  | MACC+-SD | MF1+-SD | MAUC+-SD | MMCC+-SD |
| --- | --- | --- | --- | --- |
| LCNN | 0.571+-0.058 | 0.377+-0.139 | 0.568+-0.091 | 0.112+-0.146 |
| DenseNet121 | **0.684+-0.042** | **0.466+-0.173** | **0.646+-0.068** | **0.274+-0.131** |
| ResNet50 | 0.517+-0.055 | 0.406+-0.167 | 0.584+-0.122 | 0.133+-0.174 |
| VGG16 | 0.596+-0.032 | 0.440+-0.162 | 0.610+-0.065 | 0.208+-0.134 |
| Xception | 0.569+-0.073 | 0.355+-0.136 | 0.573+-0.120 | 0.110+-0.171 |

**Figure 5:**
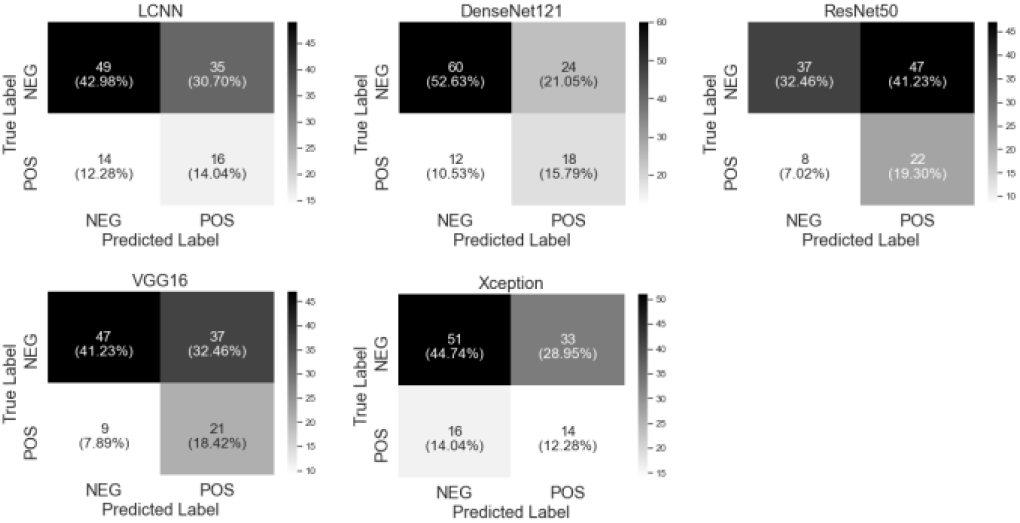
The combined confusion matrices of predicting axillary lymph node (ALN) metastases using all deep learning (DL) models during 5-fold cross-validation. LCNN is lightweight convolutional neural network.

Figure 6 reports *P*-values and average ranking between studied DL models according to Friedman post-hoc test with Bergmann and Hommel’s correction. It can be seen that DenseNet121 generates the highest average ranking among all models. The asterisk between DenseNet121 and ResNet50 means that the performance difference between the two models is significant. That is, DenseNet121 outperforms ResNet50 with statistical significance.

**Figure 6:**
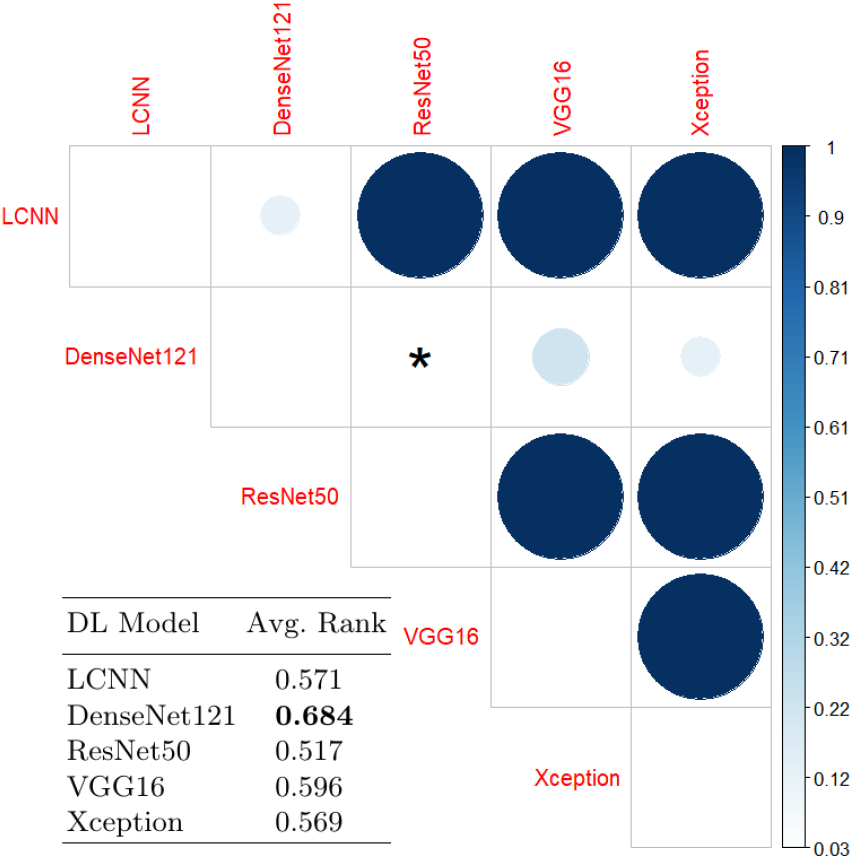
*P*-value and average ranking between all deep learning (DL) models using the Friedman test with Bergmann and Hommel’s correction. The highest average ranking (shown in bold) of a DL model indicates its performance is better than remaining models. The asterisk indicates between two DL models shows that the DL with highest ranking generates significant results as *P* < 0.05.

## IV. Discussion

Our study includes the following: (1) adapting several DL architectures including a customized convolutional neural network to address the binary classification task pertaining to detecting ALN metastases, (2) utilizing NVIDIA GeForce RTX 2080Ti GPU to evaluate DL models using 5-fold cross-validation and resize 114 gigapixel images to (256 * 256 pixels). We made these images available in supplementary material, and (3) reporting performance results using several performance measures as well as *p*-values using Friedman test with Bergmann and Hommel’s correction.

For all pre-trained models in this study, we used the feature extraction technique where we use all layers (excluding dense layers) for the feature extraction. Then, we tuned parameters of new dense layers during the training, because we changed densely connected layers to deal with the binary classification problem as in [22]. For the LCNN model, we build it from scratch with data augmentation.

For the optimization process configuration during training of DLmodels, we used RMSprop optimizer. Also, we used the binary_crossentropy loss function and the accuracy metric as in [22].

It is worth noting that DenseNet121 generated the highest results in terms of performance (Table 1) and average ranking (Figure 6) when compared to all other DL models. This coincides with recent work by Lee et al. [8], where DenseNet121 (compared against machine learning models coupled with handcrafted features) also generated the highest performance results when assessed on predicting ALN metastasis of another dataset.

## V. Conclusion and Future Work

This paper presents a DL approach for detecting ALN metastases from digital pathology images. The DL approach adapts several DL architectures, including DenseNet12, ResNet50, VGG16, Xception and a customized lightweight convolutional neural network. A training set is provided to train DL models. Then, a test set is provided to each trained model to generate predictions. We used NVIDIA GeForce RTX 2080Ti GPU to process 114 gigapixel images and evaluate models using five-fold cross-validation; these images are available in supplementary material. Experimental results show that DenseNet121 outperforms all models in this study as well as statistically significantly outperforming ResNet50.

Future work includes: (1) to further boost the performance results via incorporating clinical information combined with data augmentation techniques; and (2) to utilize other learning scenarios such as unsupervised learning for exploiting unlabeled data.

## Data Availability

All data was downloaded from public domain

## Acknowledgment

This study was supported by KAKENHI 19H05270, 20K12067, 20H04848.

