## Supplementary material for "End-to-End Deep Learning for Detecting Metastatic Breast Cancer in Axillary Lymph Node from Digital Pathology Images": ReadMe.txt

-Dataset: folder contains 114 images described as follows:
Negative axillary lymph node images: n_1.tif, n_2.tif,...,n_84.tif
Positive axillary lymph node images: p_85.tif, p_86.tif,...,p_114.tif

Please acknowledge in your publications by citing this paper and refernces at 
https://wiki.cancerimagingarchive.net/display/Public/Breast+Metastases+to+Axillary+Lymph+Nodes
